# Outcome of Wage and Self-employment intervention for Persons with Severe Mental Illness availing Rural Community-based Rehabilitation Project: Experience from South India

**DOI:** 10.1101/2024.10.31.24316167

**Authors:** Thanapal Sivakumar, Shanivaram Reddy K, Aarti Jagannathan, Channaveerachari Naveen Kumar, Jagadisha Thirthalli, Shyam K Bhat

**Affiliations:** National Institute of Mental Health and Neurosciences (NIMHANS), Bengaluru-India; The Live Love Laugh Foundation, Bengaluru, India

**Keywords:** Severe Mental Illness, Community-Based Rehabilitation, Employment, Rural health, Outcome assessment, South India

## Abstract

**Background:** The World Health Organization advocates Community-Based Rehabilitation (CBR) for resource-constrained settings. There is a need for evidence-based models of employment for persons with severe mental illness (referred to as ‘patients’) from such settings.

**Aims:** To facilitate and study the employment outcome of patients aged 18-50 years, availing a rural CBR project in South India.

**Methods:** Of 98 consented patients, only three men chose wage employment, and eighty-nine chose self-employment. Patients seeking wage employment were offered training and job placement in the nearest metropolitan city. Ten patients were offered loans for self-employment as revolving funds without collateral through the family federation of persons with mental illness. The patients and families were followed up for ten months after recruitment into intervention. The AIMS-SEEP tool assessed the impact on families that availed of loans.

**Results:** All three men who chose wage employment in the city discontinued it. Two of ten families did not use it for the intended purpose, and one loan was written off. Seven families chose sheep rearing, and one bought a tailoring machine. Self-employment was a secondary source of income for families and was used for food, clothes, school expenses, health-related costs, household items, and debt repayment. The attendance at monthly meetings of the family federation has more than tripled since loans were issued. Families reported no adverse effects due to the intervention. Only three out of eight families had repaid the loan completely at the end of ten months. Reasons cited for delay in loan repayment were hospital expenses for a sick family member and children’s school expenses.

**Conclusion:** In impoverished rural areas, patients and their families prefer self-employment locally over shifting to the city for wage employment. Suggestions for implementing livelihood interventions in other resource-constrained settings are discussed. Families must own the initiative to ensure its sustainability.

## Introduction

More than 70% of the Indian population lives in rural areas with limited awareness of mental illness and limited availability of, accessibility to, and affordability of mental health services (Gururaj et al., 2016). The World Health Organisation has advocated Community-based rehabilitation (CBR) as a feasible intervention to improve the clinical and social outcomes of persons with mental illness living in resource-constrained settings. (WHO/UNESCO/ILO/IDDC, 2010). A CBR program must address livelihood needs to ensure the sustainability of efforts. (WHO/UNESCO/ILO/IDDC, 2010).

Current evidence-based models of supported employment for persons with severe mental illness (referred to as ‘patients’ from here on) include ‘Individual placement and support’ (Bond & Mueser, 2022). They were developed in resource-rich, high-income countries with individualistic cultures and focus on ‘wage employment.’(Sivakumar & Thirthalli, 2023) India is predominantly rural, has a collectivistic culture, and offers more livelihood opportunities through self-employment. (Sivakumar & Thirthalli, 2023) In India, most patients live with their families, who address their needs and decide their treatment and employment. (Seshadri et al., 2019).

Available Indian literature on ‘employment’ is mainly from patients availing services from urban-based tertiary care centres. (Andrade et al., 2023; Haridas et al., 2021; Jagannathan et al., 2020; Jaleel et al., 2015; John et al., 2017; Khare et al., 2020, 2022; Khare, McGurk, et al., 2021; Khare, Mueser, et al., 2021). Studies from patients availing services from rural areas have described the work functioning of patients accessing treatment. (Suresh et al., 2012), income generation programs availed by treated patients with the help of their families (Ravilla et al., 2019), and various psychosocial interventions offered in the rural CBR program (Rao et al., 2022).

In rural areas, livelihood opportunities are limited, and poverty is an issue. (Suresh et al., 2012; Trani et al., 2015). Self-employment programs that tap patients’ and their families’ inherent knowledge and skills will likely be sustainable. (Rao et al., 2022; Thara et al., 2008). Many patients require family and community support to access funds, start, sustain, and succeed in self-employment. (Sivakumar & Thirthalli, 2023). There is a need to study the economic impact of livelihood support offered to patients/families in low- and middle-income countries. (Asher et al., 2017).

### Aim

To facilitate and study the employment outcome of patients aged 18-50, availing a rural CBR project in South India, according to local opportunities and family preferences.

## Material and Methods

### Setting

A rural community-based rehabilitation (CBR) program for persons with severe mental illness (referred to as ‘patients’ from here on), a partnership of the public health system with NGOs, has been implemented at [Place], a drought-prone poor taluk (subdistrict) in South India. (Sivakumar et al., 2019). Many patients on regular follow-up were started on treatment for the first time. The patients avail of psychotropic medications free of cost from the nearest government health facility and have a low level of illness severity. (Sivakumar et al., 2023).

Due to limited livelihood options, people migrate to nearby cities to earn a livelihood. Many families cannot avail themselves of bank loans for livelihood due to a lack of assets for surety, past crop loans that weren’t repaid due to drought, or ongoing loans to build a house or meet daughters’ wedding expenses. Families are reluctant to borrow from local money lenders due to the high interest rates charged.

One of the CBR program partners [Partner 1] offered employability-led domain skills training (a two-month free residential course with subsequent job placement) for wage employment and a rural livelihood entrepreneur program for self-employment of persons with disabilities.

Another CBR program partner [Partner 2], offered Rs 1,50,000 (1800 US $ at 1 US $=83.29 Indian Rupees) to the [Place] family federation to be used as a revolving fund to provide seed money to families for self-employment.

Initially, ten families would get a loan of Rs 15,000 (180 US $) each for self-employment, which had to be repaid in 10 monthly instalments (Rs 1650 (20 US $) per instalment at 1% monthly interest) at monthly meetings. The repaid money would fund fresh loans to other families for the cycle to continue.

We assured families of continued free psychiatric care, irrespective of their participation in the study. The Institutional Ethics Committee approved the study. The study was registered with the Clinical Trials Registry-India (CTRI Trial REF/2022/04/053742) dated 05^th^ May 2022.

### Participants

As of June 2022, 214 patients availing of our CBR program were approached for the study. Seventy-seven patients did not meet the inclusion criteria (28 were not residents of Taluk but were accessing the CBR program, and 49 were older than 50), and thirty-nine patients did not feel a need for employment assistance. Of the 98 patients (including 55 women) and their families who consented, only three (all men) opted for wage employment, and 89 opted for self-employment. Nine families opted out of self-employment as they were not confident about paying the monthly loan instalments.

### Intervention

#### Wage employment

The three men who opted for wage employment were referred to the state capital (approximately 240 km from [Place]) for training and job placement.

#### Self-employment

Most patients and families preferred animal husbandry for self-employment as they had the necessary expertise, space for rearing the livestock in their house, and a local market to sell it. Ten families were chosen based on resources, skills, and experience for self-employment and given a loan of Rs 15,000 (180 US $) each, which wasn’t sufficient for cow-rearing. Nine planned for sheep-rearing, and one intended to buy a tailoring machine. The families were asked to attend monthly meetings of the family federation and repay the loan in ten instalments.

The patients and families were followed up for ten months after recruitment into intervention.

### Measures

The patients who had opted for wage employment were followed up to learn the outcome of training and job placement.

The ‘Assessing the Impact of Microenterprise Services-Small Enterprise Education and Promotion (AIMS-SEEP)’ tool assessed the impact on families that availed of loans. (Nelson et al., 2005) The field staff visited families’ homes to monitor loan usage, progress and challenges faced. The attendance at monthly family federation meetings where loans were issued for self-employment and repaid was documented. The repayment rate and reasons cited for non-payment were documented.

All patients were monitored for adverse effects (including exacerbation of symptoms, relapse, hospitalisation, medication non-adherence, sleeplessness, anxiety, or accidents) due to the intervention.

## Results

### Wage employment

All three male patients who opted for wage employment returned to the village. One was uncomfortable with city life and returned without completing the training. The remaining two patients (siblings) completed training when food and accommodation were provided free of cost. High city living expenses were not commensurate with the expected salary, so they discontinued their jobs and returned to the village. One started working in a roadside restaurant for a monthly wage of Rs 7000 (85 US $). The other brother worked for local daily wages. He would earn about Rs 2000-3000 (24-36 US $) monthly from daily wages (at Rs 200 (2.4 US $) per day, assuming he gets work for 10-15 days per month). When he got an opportunity, he applied for a loan for sheep rearing from the family federation.

### Self-employment loan usage

Table 1 states the sociodemographic profile of patients who availed of loans. Two of the ten families who received the loan did not use it for the intended purpose. One family had allowed their son to spend the loan on alcohol and misled the field staff. When this was discovered by field staff, they reported an inability to repay, and the loan had to be written off. The second family repaid the loan with interest. Out of 7 families who chose sheep rearing, one had gifted the sheep kids to their married daughter’s family for their livelihood while assuming responsibility for repaying the loan. The patient and family jointly reared the sheep in the remaining six families. Two families reported the death of one sheep each due to ill health. One patient bought a tailoring machine and used it. The loan usage summary is highlighted in Figure 1.

**Table 1.** Sociodemographic profile of patients who availed of loans.

| **S. No** | **Age range (in years)** | **Sex** | **Marital Status** | **Years of Education (in years)** | **Duration of illness (in years)** | **Type of Family** | **Primary caregiver** |
| --- | --- | --- | --- | --- | --- | --- | --- |
| 1 | 40-45 | Male | Married | 12 | 24 | Extended | Wife |
| 2 | 26-30 | Female | Separated | 10 | 09 | Joint | Father |
| 3 | 26-30 | Male | Married | 10 | 20 | Joint | Father |
| 4 | 36-40 | Female | Married | 07 | 08 | Nuclear | Husband |
| 5 | 46-50 | Male | Married | 05 | 20 | Nuclear | Wife |
| 6 | 46-50 | Male | Married | 07 | 20 | Extended | Wife |
| 7 | 26-30 | Female | Married | 11 | 08 | Extended | Husband |
| 8 | 46-50 | Female | Widow | 00 | 05 | Extended | Son |
| 9 | 40-45 | Female | Married | 00 | 08 | Nuclear | Husband |
| 10 | 30-35 | Female | Married | 09 | 19 | Nuclear | Husband |

**Figure 1.**
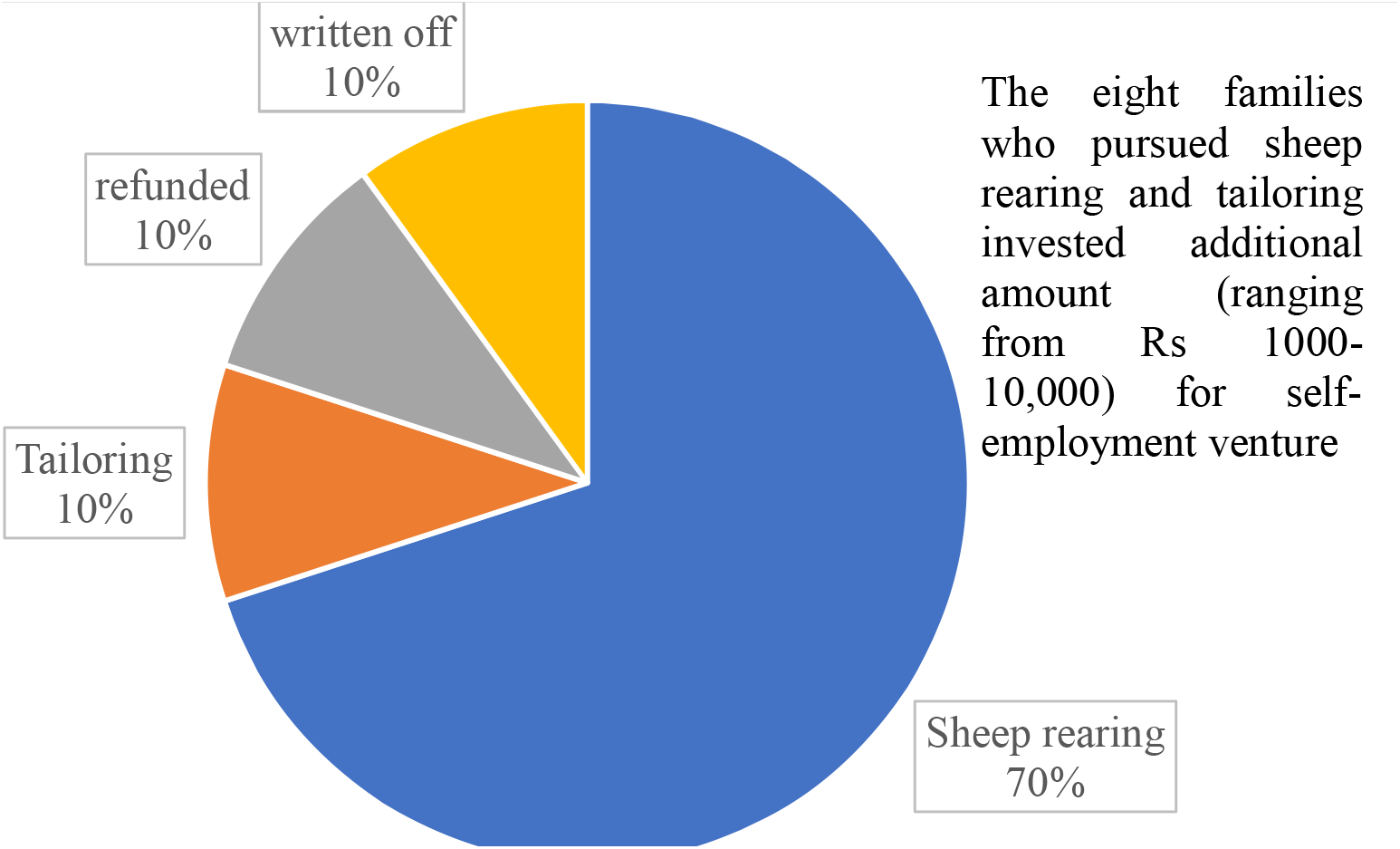
Loan usage summary.

### Regularity at follow-up consultations

All ten patients whose families had taken loans for self-employment were regular for their follow-up consultations. In comparison, out of 20 age, sex, diagnosis, and symptom-matched patients, only 12 were regular in follow-up consultations (Fisher exact test statistic 0.029).

### Attendance at monthly family federation meetings

Compared to usual attendance, attendance tripled after loans were issued.

### Repayment of loan instalments

Of the eight families, three have repaid the loan, two have 1-2 pending instalments, one has four pending instalments, and two have 6-8. Four of the eight families with pending instalments have missed 6-8 family federation meetings. The reasons cited for the delay in loan repayment were hospital expenses for a sick family member (2 families) and children’s school expenses (1 family).

### AIMS-SEEP

Three families have earned a profit/income from self-employment ranging from Rs 6000 (73 US $) monthly (tailoring machine), Rs 2000 (24 $) monthly (trading sheep monthly in the local market), and Rs 8000 (96 US $) (from the sale of 1 sheep). These families reported the profit/income as a secondary source of income and spent it on food, clothes, school expenses, health-related costs, household items, and repaying debt. They did not use it to purchase assets, expand the enterprise, or a better diet. At a ten-month follow-up, five of seven families who reared sheep were waiting for the local festive season to sell their sheep for a reasonable price. The families appreciated that the loans were offered at a lower interest than local money lenders without collateral. They felt that the loan amount was small and could be increased.

### Adverse effects of intervention

No adverse effects (including exacerbation of symptoms, relapse, hospitalisation, medication non-adherence, sleeplessness, anxiety, or accidents) were reported due to the intervention.

## Discussion

All patients who consented to the employment intervention were below the poverty line determined by the government of India. Severe mental illness was one additional disadvantage.

The families possessed skills and resources for self-employment (like knowledge of sheep rearing, space to house the sheep, availability of fodder, and local market to trade sheep). Still, they struggled to meet basic household expenses and needed money to fund self-employment. The symptomatic patient didn’t usually contribute to income among families who couldn’t afford treatment. When a low-income family has to choose between treatment for a patient or feeding the family, hunger is prioritised. They seek treatment when offered free at the nearest government health facility. (J et al., 2024). A better-resourced family spent their hard-earned money stabilising the patient’s symptoms and reported a dramatic fall in out-of-pocket expenditure when they switched to the rural CBR. (Sivakumar et al., 2019). When patients from impoverished families improve on treatment, many start working to meet their and family needs. The family and community were largely supportive.

As per local cultural norms, women do not travel far without men accompanying them, and families of female patients did not consider wage employment. Besides, families doubted the patient’s ability to care for themselves without supervision (including medication adherence) in a far-off place. All three male patients who shifted to the state capital returned to the village as they weren’t comfortable there and due to high city living expenses.

In comparison, self-employment was better accepted as patients and families could pursue it from the familiar confines of their homes. Nine families had turned down the loan as they felt they could not repay the instalments. Trust and respect for the CBR team, comprised of many people outside that locality, might have played a role. At the same time, one loan had to be written off. Starting with a lower amount to assess people’s creditworthiness and gradually raising the loan amount might help avoid such a situation. (Bandyopadhyay, 2016). Families involved the patient in sheep rearing, supervised medication adherence, and ensured basic needs were met. The patient handled the tailoring machine on her own. The families appreciated that they could repay the loan instalments at monthly meetings of the family federation, which accommodated them when they had difficulties in repayment.

Though the CBR team could ensure symptom stability through regular follow-up and facilitated a loan, the dynamics in each family were unique. One family was under financial stress due to the patient’s treatment expenses, which stabilised after CBR. The patient’s spouse was resourceful. He used the loan well to generate a monthly income, surprising the CBR staff. Two families with regular income incurred problems due to alcohol use (the patient himself) and gambling (by the patient’s spouse). Repaying the loan was not the top priority as they had to manage household expenses (including children’s education) with available money.

After discussion, the family federation offered loans for purposes other than self-employment. From the repaid loan instalments, 12 fresh loans have been issued to new beneficiaries for livelihood (3 for sheep rearing, 1 for cow rearing, 1 for vegetable selling, 1 to improve petty shop, 2 for agricultural activities), children’s education (2 families) and household expenses (2 families).

The fine charged for default in loan repayment was Rs 150 (1.2 US $) for three consecutive defaults for the initial ten loans issued. We found that some families would attend the family federation meeting only on the third month and pay one instalment to avoid the fine. The fine was revised to Rs 50 (0.6 US $) for each default for these loans. This has ensured better attendance at the monthly family federation meeting and repayment rate for the 12 fresh loans.

Poverty is multifactorial and complex and requires multipronged interventions. (Banerjee & Duflo, 2011; Lund et al., 2011). Livelihood interventions offered as part of CBR for patients from impoverished rural areas will be a small step in that direction. Based on our experience, the following suggestions can be considered for implementing similar initiatives in other resource-constrained settings.

- Organisations offering community treatment for patients in rural areas can establish a family federation comprising patients and their caregivers.
- A proactive family member must be identified to lead the family federation.
- The family federation can be encouraged to conduct at least monthly meetings at the local government health facility (usually well connected by road) with the prior permission of concerned officials.
- During the meeting, families must be encouraged to share their personal experiences caring for the patient. The families can be educated about various avenues (including government schemes) for empowering the patient. Discussion on challenges faced by families needs to be facilitated. Families need to advocate for change in the system to meet their needs. This will facilitate cohesiveness among the group.
- Funds for the family federation can be generated from yearly membership fees collected from each family, donors, or bank loans.
- The members of the family federation can be offered loans at lower-than-market interest rates for self-employment or other household expenses (like children’s education or repairing houses). This is especially helpful for families of less skilled patients from agrarian communities.
- Family members who regularly attend the family federation meetings (at least three consecutive meetings) can be considered for issuing loans.
- Before issuing the loan, a home visit is needed to assess the family living conditions, family dynamics, nature of the relationship between the patient and other family members, who are the primary caregivers, whom the patient listens to, available resources for self-employment, and how the family will repay the loan amount.
- It is desirable to start with a small initial loan amount. If the family repays successfully, increase the loan amount gradually. This helps identify ‘credit-worthy’ families who may benefit from higher loans.
- The loan agreement with a family member should specify the following terms and conditions: total loan amount, number of instalments, repayment amount per instalment, interest amount per instalment, fine for default in repaying instalments, responsibility of family to use funds responsibly, that the loan is issued by family federation in good faith and federation is not responsible if the family faces challenges in the venture for which loan was given, and family needs to repay irrespective of the outcome of the venture.
- A monthly fine for default in loan repayment needs to be charged.
- Periodic home visits are required to ensure the loan amount is used for the intended purpose.
- Families may face genuine hardships (like the breadwinner of a family falling sick). Home visits can corroborate this. In such cases, the family federation must be considerate and permit delayed repayment.
- Families with at least two earning members have better chances of repaying the loan amount.
- Families where one family member (usually male) regularly consumes alcohol or gambles find it challenging to meet family expenses and repay the loan instalments. This is a red flag to be watched for.

## Conclusion

In impoverished rural areas, patients and their families prefer self-employment locally rather than shifting to the city for wage employment. Apart from ensuring community mental health services, the Government should fund self-employment livelihood options for low-income families of patients in rural areas through revolving funds. Only family members caring for the person with mental illness should be issued loans. Such families feel rewarded for their caregiving efforts and perceive the patient positively for helping them access precious capital for a livelihood venture. Apart from reducing economic burden, self-employment allows families to engage patients in gainful work. It also promotes patients’ dignity and social inclusion in the local community. Families need to own the initiative to ensure its sustainability.

## Data Availability

All data produced in the present study are available upon reasonable request to the authors

## Acknowledgements

The authors thank the Department of Psychiatry, NIMHANS (Bengaluru), National Health Mission (Bengaluru), District Health Officer (Davangere), District Leprosy Officer (Davangere), District Mental Health Program (Davangere), Taluk Health Officer (Jagaluru), all Jagaluru taluk health care staff, patients, their family caregivers, and officials/ key community members from Jagaluru Taluk for their support and cooperation.

The work was made possible with the support of the Association of People with Disability: [Mr Janardhana AL (Director: Mental Health Program & Life Cycle Approach), Mr Santosha S (Program Manager, Community mental health program)], Chittasanjeevini charitable trust, and Mr Dundappa Doddur (project staff).

The authors thank The Live Love Laugh Foundation (TLLLF) staff (Ms Anisha Padukone, Ms Kainaat Khan and Ms Ankita Mathews) for their advice on study design, interpretation, and helpful comments on manuscript drafts.

